# LLIN Evaluation in Uganda Project (LLINEUP2) – Effect of long-lasting insecticidal nets (LLINs) treated with pyrethroid plus pyriproxyfen vs LLINs treated with pyrethroid plus piperonyl butoxide in Uganda: a cluster-randomised trial

**DOI:** 10.1101/2024.07.31.24311272

**Authors:** Samuel Gonahasa, Jane F Namuganga, Martha J Nassali, Catherine Maiteki-Sebuguzi, Isaiah Nabende, Adrienne Epstein, Katherine Snyman, Joaniter I Nankabirwa, Jimmy Opigo, Martin J Donnelly, Grant Dorsey, Moses R Kamya, Sarah G Staedke

## Abstract

**Background:** We embedded a pragmatic, cluster-randomised trial into Uganda’s national campaign to distribute long-lasting insecticidal nets (LLINs) in 2020–2021, comparing LLINs treated with pyrethroids plus the synergist piperonyl butoxide (PBO), to LLINs treated with pyrethroids plus pyriproxyfen, an insect growth regulator.

**Methods:** Target communities surrounding public health facilities (clusters, n=64), covering 32 high-burden districts not receiving indoor residual spraying, were included. Clusters were randomised 1:1 in blocks of two by district to receive: (1) pyrethroid-PBO LLINs (PermaNet 3.0, n=32) or (2) pyrethroid-pyriproxyfen LLINs (Royal Guard, n=32). LLINs were delivered from 7 November 2020 to 26 March 2021 and malaria outcome data were collected until 31 March 2023. Estimates of malaria incidence in residents of all ages (the primary outcome) were generated for each cluster from enhanced health facility surveillance data. At 12- (24 November 2021 to 1 April 2022) and 24-months (23 November 2022 to 21 March 2023) post-LLIN distribution, cross-sectional community surveys were conducted in randomly selected households (at least 50 per cluster, 3,200 per survey).

**Findings:** In the two years following LLIN distribution, 186,364 episodes of malaria were diagnosed in cluster residents during 398,931 person-years of follow-up. Malaria incidence after 24-months was lower than at baseline in both arms (pyrethroid-PBO: 465 vs 676 episodes per 1000 person-years; pyrethroid-pyriproxyfen: 469 vs 674 episodes per 1000 person-years); but the difference between the arms was not statistically significant (incidence rate ratio 1.06, 95% confidence interval [CI] 0.91– 1.22, p=0.47). Two years post-distribution, ownership of at least one LLIN for every two household residents was low in both arms (41.1% pyrethroid-PBO vs 38.6% pyrethroid-pyriproxyfen). Parasite prevalence in children aged 2-10 years was no different between the arms in either survey and similar results were observed for prevalence of anaemia in children aged 2-4 years.

**Interpretation:** The effectiveness of pyrethroid-PBO LLINs and pyrethroid-pyriproxyfen LLINs was no different in Uganda, but two years after mass distribution, LLIN coverage was inadequate.

**Funding:** US National Institutes for Health, Bill and Melinda Gates Foundation

**Trial registration:** NCT04566510. Registered 28 September 2020, https://clinicaltrials.gov/ct2/show/NCT04566510

## Introduction

Between 2000 and 2015, intensified efforts to control malaria substantially reduced malaria case incidence and parasite prevalence across Africa, with the majority of cases averted attributed to the distribution of long-lasting insecticidal nets (LLINs) [1]. More recently, however, progress on malaria control has stalled. Globally, malaria cases increased from 230 million in 2015 to 247 million in 2021, with 95% of cases reported from Africa [2]. Four African countries, including Uganda, accounted for nearly half of all cases worldwide in 2021. In Uganda, implementation of malaria control interventions including repeated national LLIN distribution campaigns, case management with effective artemisinin-based combination therapy, and indoor residual spraying (IRS) of insecticides in selected high-transmission districts reduced the malaria burden, with parasite prevalence by microscopy in children under-five decreasing from 45% in 2009 to 9% in 2018-2019 [3]. Progress in malaria control in Uganda has been difficult to sustain, however, despite ongoing implementation of effective control interventions [4, 5].

In Uganda, and across Africa, LLINs are the most widely-used malaria prevention tool [6]. As recommended by the World Health Organization (WHO), the Ugandan Ministry of Health distributes LLINs through mass campaigns approximately every three years, aiming for universal coverage [7]. Currently, LLINs rely on pyrethroid insecticides, but resistance to pyrethroids has spread rapidly in malaria vectors, threatening the effectiveness of LLINs [2, 8]. The causal mechanisms of pyrethroid resistance are complex, but point mutations in the voltage-gated sodium channel where pyrethroids bind (‘knock down resistance’), and alterations in enzymes that metabolize pyrethroids, commonly cytochrome P450s, are key features [9, 10].

To combat emerging resistance, next generation LLINs that incorporate a second chemical in addition to pyrethroids have been developed. One such net includes piperonyl butoxide (PBO), a synergist which inhibits P450 enzymes, overcoming metabolic resistance mechanisms in mosquitoes, and partially restoring susceptibility to pyrethroids [11, 12]. Supported by evidence from trials conducted in Tanzania and Uganda [13–15], demonstrating the superiority of pyrethroid-PBO LLINs over pyrethroid-only LLINs, the WHO endorsed deployment of pyrethroid-PBO LLINs in areas where malaria vectors exhibit pyrethroid resistance. The WHO’s recommendation was conditional due to uncertainty about the cost-effectiveness and durability of pyrethroid-PBO LLINs [16, 17]. Another new type of LLIN combines pyrethroids with pyriproxyfen, an insect growth regulator which acts as a sterilising agent, reducing the vector population density and lifespan of adult mosquitoes. Pyrethroid-pyriproxyfen LLINs are promising, but trials comparing these nets to pyrethroid-only LLINs in Burkina Faso and Tanzania have produced mixed results [18, 19]. Additional studies evaluating the effectiveness of nets incorporating pyriproxyfen are needed.

In 2020-2021, two next generation LLINs including pyrethroid-PBO LLINs and pyrethroid-pyriproxyfen LLINs were distributed in Uganda, providing an opportunity to evaluate the effectiveness of these LLINs in a ‘real-world’ setting. With support from the Ministry of Health, donors, and partners, a large cluster-randomised trial (LLINEUP2) was embedded within this national distribution campaign, allowing us to rigorously compare the impact of these LLINs on epidemiological indicators. We aimed to test the hypothesis that malaria incidence would be lower in intervention clusters (randomised to receive pyrethroid-pyriproxyfen LLINs) than in control clusters (randomised to receive pyrethroid-PBO LLINs). Malaria surveillance data at health facilities were collected between 17 October 2020 and 31 March 2023, and cross-sectional community survey data between 24 November 2021 to 1 April 2022 (12-months) and 23 November 2022 to 21 March 2023 (24-months) post-LLIN distribution.

## Methods

### Study setting

Districts with high malaria burden were selected to participate in the study (figure 1) if they were not receiving IRS and were scheduled to receive pyrethroid-PBO LLINs. A cluster was defined as the target area surrounding public health facilities with enhanced malaria surveillance, known as Malaria Reference Centres (MRCs) [20]. MRCs are high-volume, level III or IV health centres with an estimated outpatient department attendance of 1,000–3,000 patients per month and a functional laboratory. To avoid contamination, MRCs within the same district were selected from different sub-counties. Overall, 64 clusters located in 32 districts were included in the trial.

**Figure 1:**
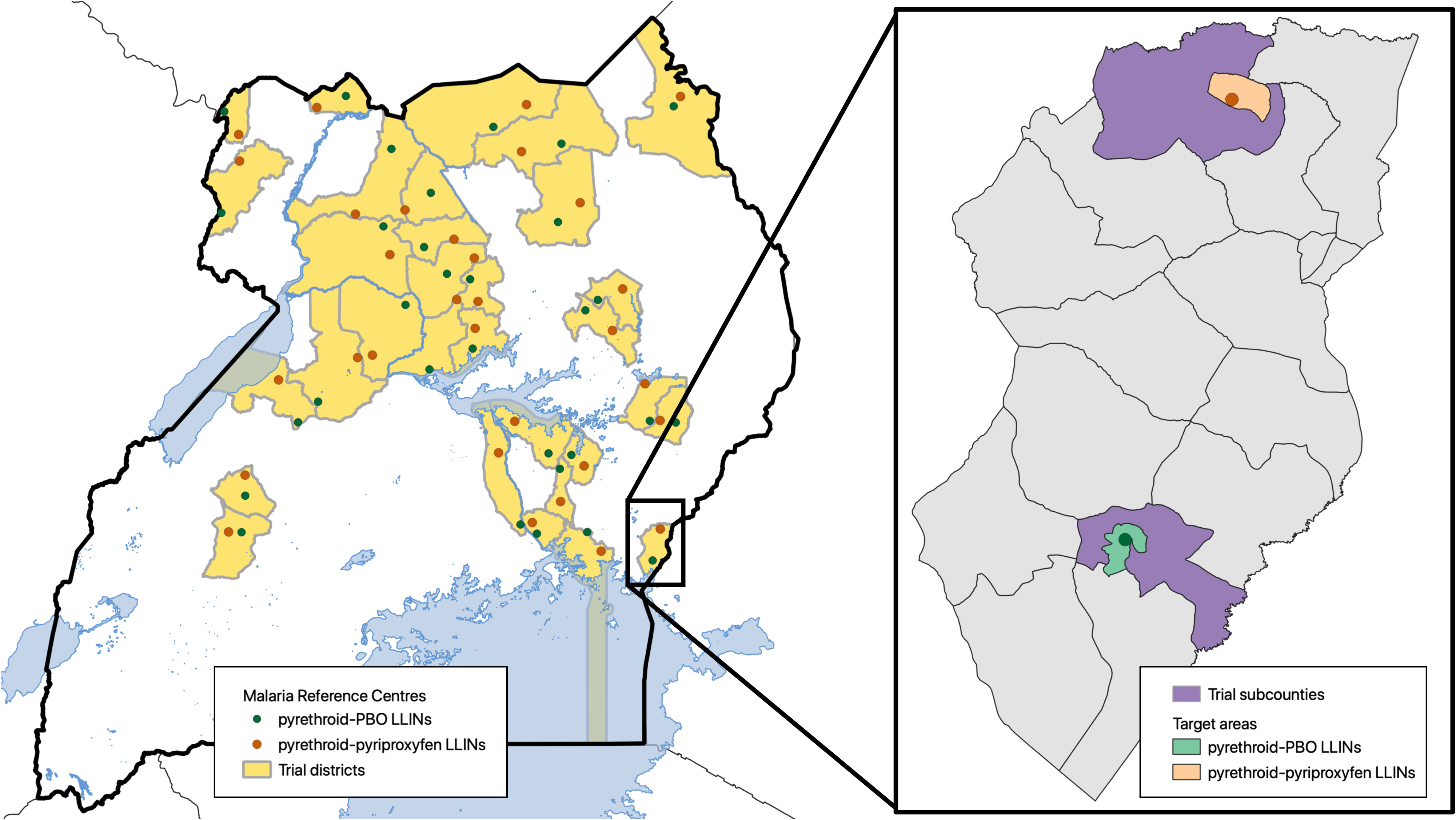
Map of study area: The study included 64 clusters across 32 districts; 2 clusters per district, one per study arm. A cluster was defined as a malaria reference centre and its target area.

### Assignment of interventions

#### Cluster definition

The unit of randomisation (cluster) was the ‘target area’ surrounding each MRC. Target areas had an estimated population of at least 1,200 residents and included the MRC’s village plus adjacent villages without another health facility, located within the same sub-county as the MRC village, and with similar malaria incidence as the MRC village. All households within the target areas were mapped and enumerated to determine the population and to generate a sampling frame for community surveys.

#### Cluster randomisation

Clusters were randomised in a 1:1 ratio to receive either pyrethroid-PBO LLINs (PermaNet 3.0, Vestergaard Sàrl, Switzerland), or pyrethroid-pyriproxyfen LLINs (Royal Guard, Disease Control Technologies, USA). Randomisation was carried out by a member of the study team based outside of Uganda at the University of California, San Francisco, who had no direct links to the net distribution team at the Ministry of Health in Uganda. To improve comparability between arms, MRCs were selected in each district for allocation to the intervention and control arm in a matched pair-wise fashion. Clusters were randomised in blocks of two, with each block representing a district containing two clusters, using the ‘runiform’ command in STATA version 14.2 (StataCorp, Texas, USA).

#### LLIN distribution

LLINs were delivered free-of-charge through a mass distribution campaign led by the Ugandan Ministry of Health and partners from 7 November 2020 to 26 March 2021. Members of the research team engaged with Uganda’s national committees that coordinated the campaign, to ensure that LLINs were allocated according to the randomisation scheme. For each cluster we used a ‘fried egg’ approach (figure 1); LLINs were delivered to the entire sub-county, including the MRC and several other villages beyond the MRC target area (‘egg white’), while outcomes were measured in the MRC target areas (‘egg yolk’). Nets were distributed from house-to-house, targeting all households within the subcounty and aiming for population level coverage of at least 85%.

### Health facility-based surveillance

At each MRC, individual-level data for all patients recorded into standardised outpatient department registers were entered into an Access database by on-site data entry officers. Primary data captured included location of residence (parish and village), age, gender, temperature, history of fever, type of malaria test done, malaria test results, diagnoses given, and any treatments prescribed. Data from each MRC was submitted to the research team monthly using a secure on-line system. Data captured from 1 November 2019 to 31 March 2023 was used to generate estimates of malaria incidence.

### Cross-sectional surveys

Cross-sectional community surveys were conducted 12-months (24 November 2021 to 1 April 2022) and 24-months (23 November 2022 to 21 March 2023) after nets were distributed. A study team, consisting of a clinician and research assistant carried out the surveys. Households were randomly selected from the enumeration lists for each cluster and screened until 50 households with at least one child aged 2–10 years was enrolled from each cluster (with a total of 64 clusters, leading to at least 3,200 households per survey). If an enrolled household had no children of appropriate age, they were included in the household survey only, but did not take part in the clinical survey. Households were included if at least one adult aged 18 years or older was present, the adult was usually resident and slept in the household on the night before the survey, and the adult provided informed consent. Households were excluded if the house had been destroyed or could not be found, the house was vacant, or no adult resident was home on more than three occasions.

A household questionnaire adapted from prior surveys, including the national Malaria Indicator Survey, was administered to the head of the household (or their designate), after obtaining their consent, using a hand-held tablet computer. Information was gathered on the characteristics of households and residents, proxy indicators of wealth including ownership of assets, and ownership and use of LLINs in the households.

All children aged 2–10 years from enrolled households were eligible for the clinical survey if they were usually resident and present in the household on the night before the survey, their parent or guardian provided informed consent, and the child provided assent if they were aged 8 years or older. Children not home on day of survey were excluded. The clinical surveys included measurement of temperature, subjective fever, and a finger-prick blood sample for preparation of thick blood smears, and haemoglobin measurement (in children aged 2–4 years).

Participants with a temperature of ≥38.0°C, or who reported fever in the past 48h, had a rapid diagnostic test (RDT) done (Bioline Malaria Ag P.f/Pan, Abbott Diagnostics, Inc). Participants with a positive RDT were treated with artemether-lumefantrine, or referred if they had evidence of severe disease. Any participants with a haemoglobin <7.0 g/dL or other concerning clinical symptoms were referred for further management.

### Laboratory procedures

Thick blood smears obtained in the cross-sectional surveys were dried and transported to the Infectious Diseases Research Collaboration Molecular Research Laboratory in Kampala within seven days for processing and reading. Slides were stained with 3% Giemsa for 30 minutes and read by experienced laboratory technologists. Parasite densities were calculated by counting the number of asexual parasites, per 200 leukocytes (or per 500, if the count was less than 10 parasites per 200 leukocytes), assuming a leukocyte count of 8,000/μl. A thick blood smear was considered negative when the examination of 100 high power fields did not reveal asexual parasites. For quality control, all slides were read by a second microscopist and a third reviewer settled discrepant readings, defined as (1) positive versus a negative thick blood smear, (2) parasite density differing by ≥25%. Haemoglobin measurements were made using a portable HemoCue analyzer (HemoCue, Ängelholm, Sweden).

### Outcome measures

The primary outcome of the trial was cumulative incidence of symptomatic malaria cases in cluster residents of all ages captured at the MRCs over the 24 months following LLIN distribution. The total number of malaria cases for each cluster was determined from the number of laboratory-confirmed malaria cases diagnosed at the MRC in patients residing within the target area, over the two-year follow-up period, correcting for cluster residents with suspected malaria seen at the MRC who did not undergo laboratory testing, and for patients with laboratory-confirmed malaria whose village of residence was unknown (due to missing data). The population of the MRC target area determined in the enumeration surveys was used as the denominator for incidence calculations for each cluster with a constant growth factor of 0.29% per month, derived from the UN Department of Economic and Social Affairs Population Division World Population Prospects data (https://population.un.org/wpp/), which estimated an annual growth rate of 3.46% in 2019 (divided by 12 for the monthly rate of 0.29%).

Secondary outcomes for the cross-sectional community surveys included LLIN ownership (the proportion of households that owned at least one LLIN), adequate LLIN coverage (the proportion of households that owned at least one LLIN for every two occupants), LLIN use (the proportion of household residents who reported sleeping under an LLIN the previous night), prevalence of parasitaemia (in children aged 2–10 years), and prevalence of anaemia (in children aged 2–4 years).

### Sample size calculations

Our sample size of 32 clusters per arm was calculated to detect a 26% difference (incidence rate ratio [IRR] = 0.74) in malaria incidence between the two study arms over the 24 months following LLIN distribution (the primary endpoint of the study), given a power of 80% and a two-sided significance level of 0.05, assuming an incidence of 332 malaria cases per 1,000 person-years in the control arm and a coefficient of variation of 0.42, which was calculated from the 14 MRCs where estimates of malaria incidence over 6 months were available at the time of protocol development.

For the cross-sectional community surveys, we planned to sample all eligible children aged 2–10 years from 50 households in the 64 clusters in each survey. Assuming an average of 1.8 children aged 2–10 years per household, we estimated that 5,760 children from 3,200 households would be included. Assuming a coefficient of variation of 0.40, across a wide range of prevalence measures in the control arm (10–70%), our sample size allowed us to detect a 25–28% difference in the prevalence measure of interest, equivalent to a prevalence ratio of 0.72–0.75, given a power of 80% and a two-sided significance level of 0.05.

### Analytical issues

All analyses were conducted using an intention-to-treat approach. We compared cluster-level estimates of the incidence of malaria between the intervention (pyrethroid-pyriproxyfen LLINs) and the control arm (pyrethroid-PBO LLINs) using a mixed effects Poisson regression model with a random effect at the district level (each district included two clusters, randomised to one of the two study arms) and adjustment for log-transformed estimates of the baseline malaria incidence (three months prior to the intervention) for each cluster. The effect of the intervention was expressed as an incidence rate ratio (incidence in the intervention arm/incidence in the control arm).

For the cross-sectional surveys, separate analyses were conducted 12- and 24-month following LLIN distribution. Houses were classified as ‘modern’ (synthetic walls and roofs, eaves closed or absent) or ‘traditional’ (all other construction). We compared individual level estimates of the prevalence of parasitaemia and anaemia between the intervention (pyrethroid-pyriproxyfen LLINs) and the control arm (pyrethroid-PBO LLINs) using a mixed effects logistic regression model with random effects at the level of the cluster and household. We compared household-level estimates of LLIN ownership, coverage, and use with a mixed effects logistic regression model with a random effect at the level of the cluster. Effects of the intervention were expressed as odds ratios (odds in the intervention arm/odds in the control arm).

### Research ethics approval and participant consent

The research included in this trial was conducted under two study protocols. The Implementation Project Pilot Study covered enumeration of the target areas and estimation of malaria incidence using health facility surveillance data. This project was approved by the Makerere University School of Medicine Research & Ethics Committee (SOMREC, Ref 2019-087), the Ugandan National Council of Science and Technology (UNCST, Ref HS 2659), and the University of California, San Franciso Human Research Protection Program Institutional Review Board (UCSF Ref 19-27957).

The LLINEUP2 trial protocol provided approval for the full trial, including the cross-sectional surveys, and was approved by the Makerere University School of Medicine Research & Ethics Committee (SOMREC Ref 2020-193), Ugandan National Council for Science and Technology (UNCST Ref HS1097ES), the University of California San Francisco Human Research Protection Program Institutional Review Board (UCSF Ref 20-31769), and the London School of Hygiene & Tropical Medicine Ethics Committee (LSHTM Ref 22615). Prior to enrolment into the clinical surveys, written informed consent was obtained from a parent or guardian for all children aged less than 18 years and the child provided assent if they were aged 8 years or older.

## Results

### Characteristics of clusters, households, and residents

Of the 64 clusters (figure 2), 32 were randomised to receive pyrethroid-PBO LLINs, and 32 to receive pyrethroid-pyriproxyfen LLINs. At baseline, the median estimated population of the clusters (MRC target areas) was 3,070 (range 1,084—5,915) in the pyrethroid-PBO LLIN arm, and 2,493 (range 756—8,663) in the pyrethroid-pyriproxyfen LLIN arm.

**Figure 2:**
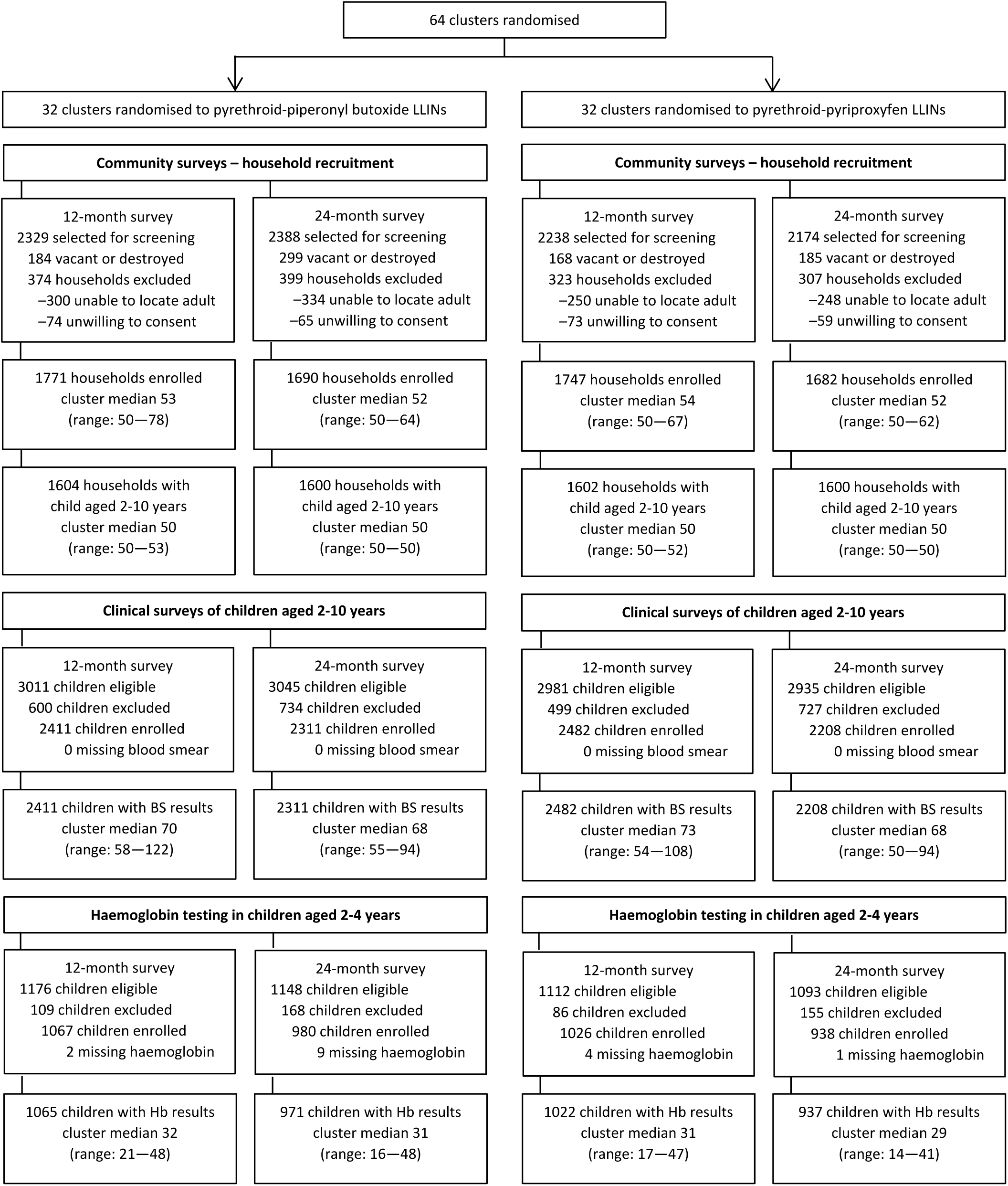
Trial profile. LLIN=long-lasting insecticidal net. BS=blood smear. Hb=haemoglobin.

Cross-sectional community surveys were carried out in all clusters (table 1) at 12-months (24 November 2021 to 1 April 2022) and 24-months (23 November 2022 to 21 March 2023) post-LLIN distribution. Characteristics of households enrolled in both surveys were similar across study arms (12-months: n=3,518, median 53 per cluster, range 50⍰78; 24-months: n=3,372, median 52 per cluster, range 50⍰64). In both surveys, over half of households had three or more persons per sleeping room (55.8%) and were constructed of traditional materials (57.1%). Overall, 7,781 bed nets were surveyed at 12-months, and 6,306 at 24-months (table 1). Most nets (86.1%) were obtained in the most recent universal coverage campaign (UCC), and nearly all nets (96.1%) were obtained for free.

**Table 1.**
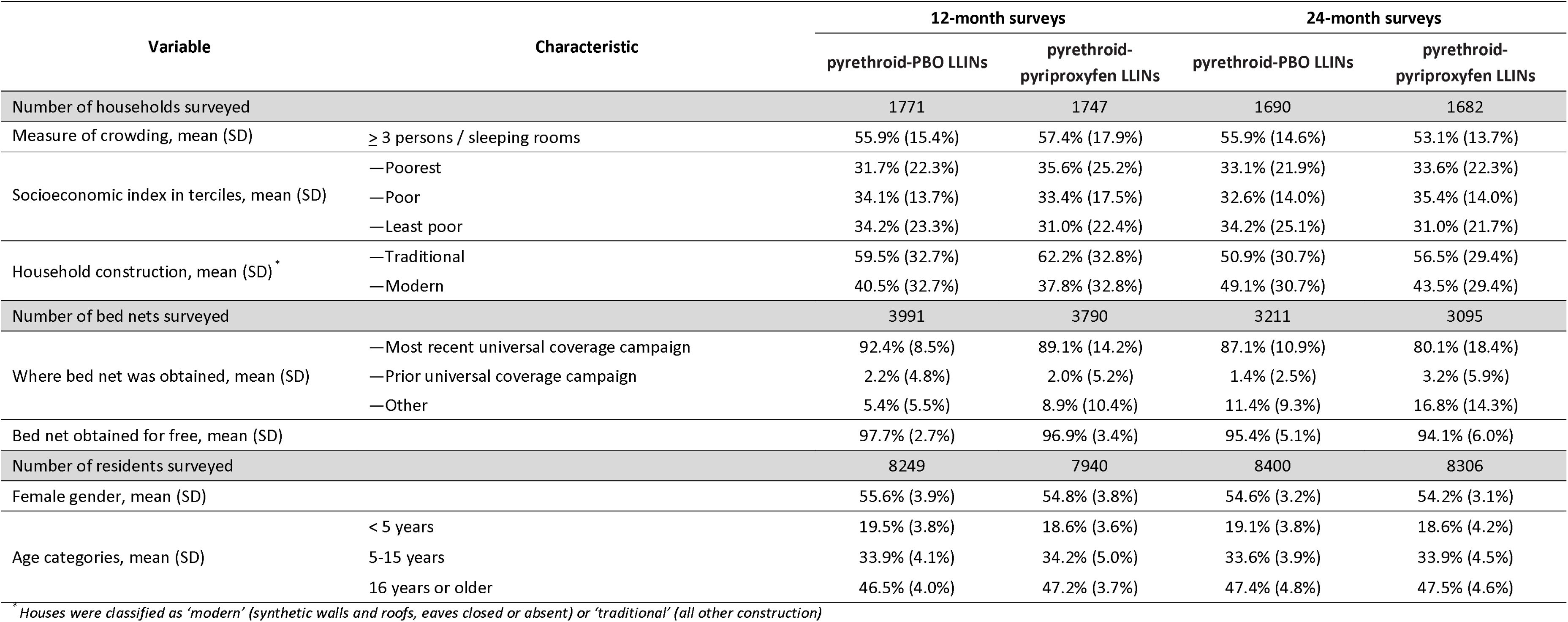
Cluster-level characteristics of households, bed nets and household residents from cross-sectional surveys.

In total, 16,189 household residents were surveyed at 12-months, and 16,706 at 24-months (table 1); over half of residents were women (54.5%). In both surveys, the age distribution of residents was similar across the study arms. Children aged 2-10 years enrolled in the clinical survey (figure 2) were tested for parasitaemia (12-months: n=4,893, median 73 per cluster, range 541⍰22; 24-months: n=4,519, median 68 per cluster, range 50⍰94). Children aged 2-4 years were tested for anaemia (12-months: n=2,087, median 31 per cluster, range 17⍰48; 24-months: n=1,908, median 30 per cluster, range 14⍰48).

### LLIN delivery

LLINs were delivered in the study area by the Ministry of Health and partners from 7^th^ November 2020 to 26^th^ March 2021. In total, 1,329,273 LLINs were allocated for distribution to the sub-counties surrounding each cluster, including 632,359 pyrethroid-pyriproxyfen LLINs and 696,914 pyrethroid-PBO LLINs. A cross-sectional survey was conducted in April-May 2021 in 12 clusters from 12 selected districts to evaluate the impact of the LLIN distribution campaign. As previously reported,[21] 1-5 months after LLIN distribution, household LLIN ownership was high. Of 634 households surveyed, 609 (96.1%) owned at least one LLIN. However, only 360 (56.8%) households owned at least one UCC LLIN for every 2 residents, while 374 (59.0%) were adequately covered by LLINs of any type.

### Impact on primary outcome: malaria incidence

In the three months prior to completion of LLIN distribution (baseline), 32,384 episodes of malaria were diagnosed in residents of the clusters (MRC target areas) over 47,945 per-years of follow-up (table 2, figure 3). The baseline incidence of malaria in the two study arms was similar (676 episodes per 1,000 person-years in the pyrethroid-PBO LLIN arm, and 674 episodes per person-year in the pyrethroid-pyriproxyfen LLIN arm). In the first 4 months following LLIN distribution, malaria incidence decreased in both study arms, but began to increase again 5-months post-distribution (figure 3). Overall, in the two years after LLIN distribution, 186,364 episodes of malaria were diagnosed in cluster residents during 398,931 person-years of follow-up (table 2). The incidence of malaria post-LLIN distribution was lower than baseline in both study arms (465 episodes per 1000 person-years in the pyrethroid-PBO arm, and 469 episodes per person-year in the pyrethroid-pyriproxyfen arm), but the difference between the two arms was not statistically significant (incidence rate ratio (IRR) 1.06, 95% confidence interval (CI) 0.91–1.22, p⍰=⍰0.47).

**Figure 3:**
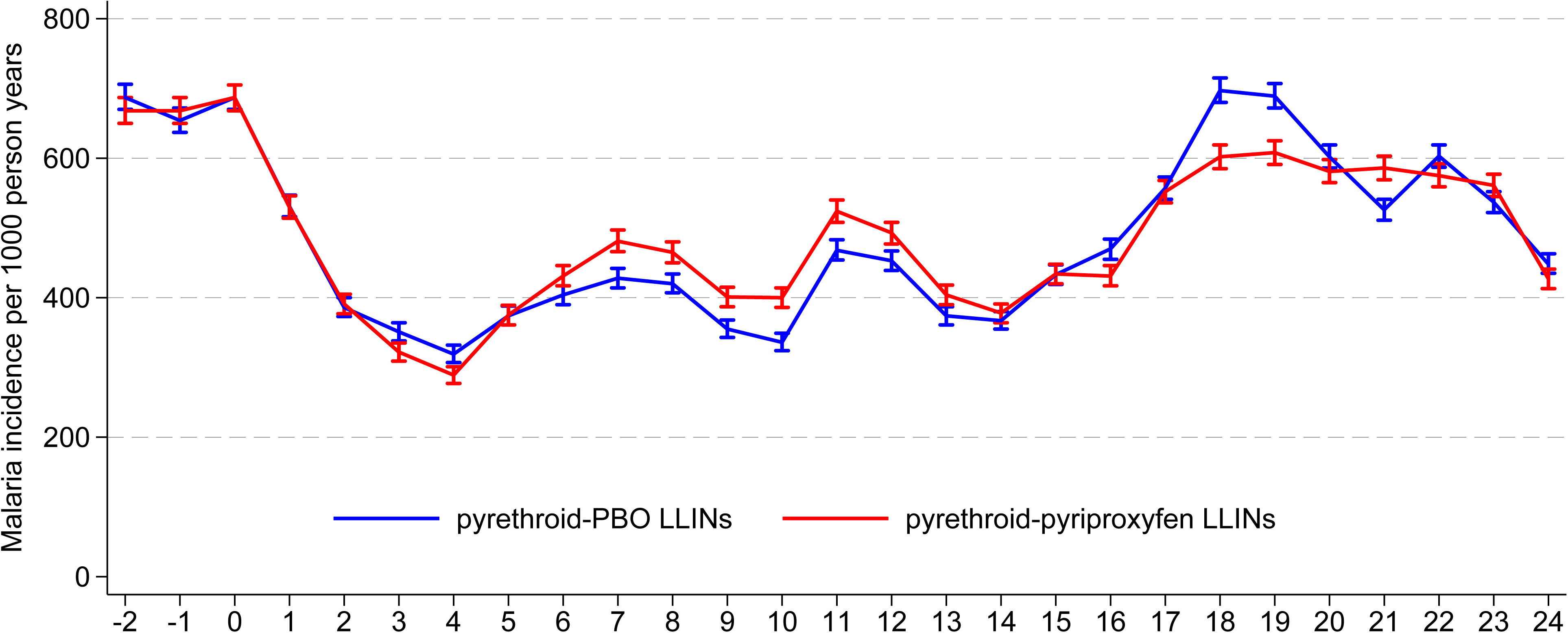
Malaria incidence per 1000 person-years. LLINs were distributed in waves over a period of 5 months. Malaria incidence measures cater for the staggered distribution of LLINs across the sites.

**Table 2.** Malaria incidence in residence of all ages from MRC target areas.

| Study arm | Baseline * |  |  | 24 months post-intervention |  |  | IRR (95% CI)** | p-value |
| --- | --- | --- | --- | --- | --- | --- | --- | --- |
|  | Episodes of malaria | Person years of observation | Malaria incidence per 1000 PY | Episodes of malaria | Person years of observation | Malaria incidence per 1000 PY |  |  |
| pyrethroid-PBO LLINs | 16,763 | 24,783 | 676 | 95,898 | 206,211 | 465 | reference arm |  |
| pyrethroid-pyriproxyfen LLINs | 15,621 | 23,162 | 674 | 90,466 | 192,720 | 469 | 1.06 (0.91-1.22) | 0.47 |

\* 3 months prior to completion of LLIN distribution
\*\* Adjusted for baseline malaria incidence; random effect at the level of the district

### Impact on secondary outcomes: LLIN ownership, coverage and use

Trends in LLIN ownership, coverage and use were similar in both study arms at 12-months and 24-months (table 3). Most households reported owning at least one LLIN at 12-months (91.9% pyrethroid-PBO vs 89.9% pyrethroid-pyriproxyfen), but ownership dropped at 24-months in both study arms (82.5% pyrethroid-PBO vs 80.0% pyrethroid-pyriproxyfen). In contrast, adequate coverage of LLINs (one LLIN for every two residents) was low at 12-months (59.9% pyrethroid-PBO vs 56.6% pyrethroid-pyriproxyfen) and fell even further at 24-months (41.1% pyrethroid-PBO vs 38.6% pyrethroid-pyriproxyfen). Similarly, although most household residents reported sleeping under a LLIN the previous night (74.2% pyrethroid-PBO vs 76.1% pyrethroid-pyriproxyfen); reported use of LLINs dropped at 24-months (63.6% pyrethroid-PBO vs 61.6% pyrethroid-pyriproxyfen).

**Table 3.**
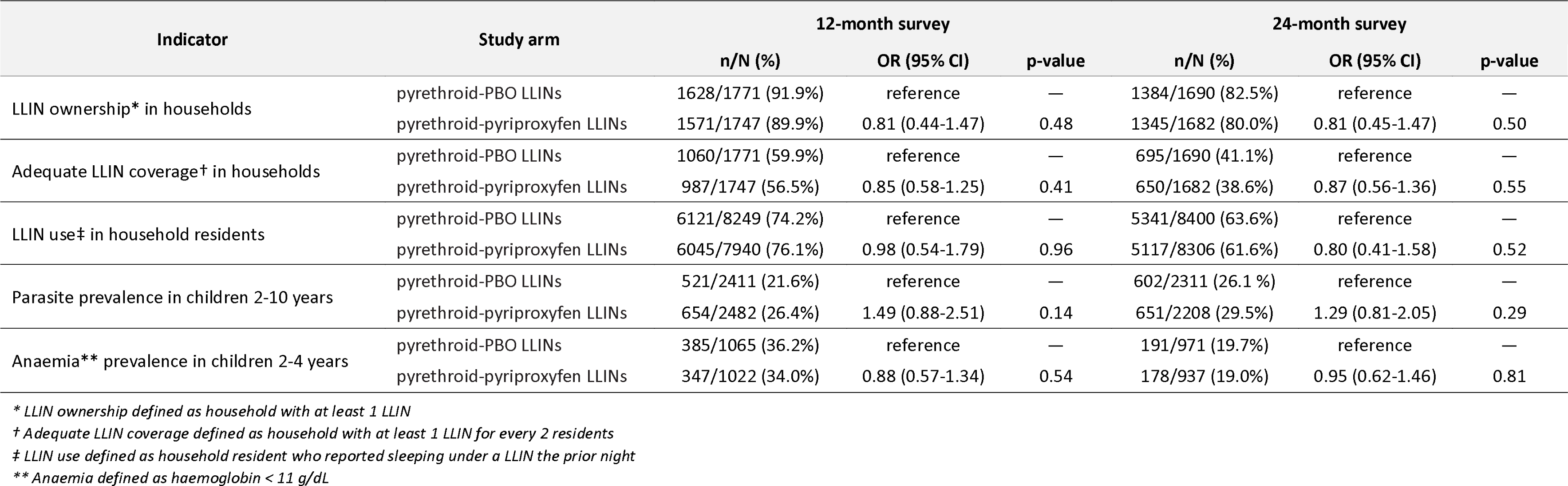
Secondary outcomes assessed at 12- and 24-months post-intervention from cross-sectional surveys of MRC target areas.

### Impact on secondary outcomes: parasite prevalence and anaemia prevalence

Parasite prevalence in children aged 2-10 years was no different in the two study arms at 12-months and 24-months post-LLIN distribution (table 3). Parasite prevalence at 12-months was 21.6% in the pyrethroid-PBO arm vs 26.4% in the pyrethroid-pyriproxyfen arm (odds ratio [OR] 1.49 [95% CI: 0.88–2.51], p=0.14). Similar results were observed at the 24-month timepoint (26.1% in the pyrethroid-PBO arm vs 29.5% in the pyrethroid-pyriproxyfen arm (OR 1.29 [95% CI: 0.81–2.05], p=0.29). The prevalence of anaemia (haemoglobin < 11 g/dL) in children aged 2-4 years decreased substantially between 12-months and 24-months, but there was no difference between the study arms at the two timepoints (table 3). Anaemia prevalence at 12-months was 36.2% in the pyrethroid-PBO arm vs 34.0% in the pyrethroid-pyriproxyfen arm (OR 0.88 [95% CI: 0.57–1.34], p=0.44). At 24-months, prevalence anaemia dropped to 19.7% in the pyrethroid-PBO arm vs 19.0% in the pyrethroid-pyriproxyfen arm (OR 0.95 [95% CI: 0.62–1.46], p=0.81).

### Costing of LLINs

We originally planned to conduct a cost-effectiveness analysis of the two LLINs studied, but given the lack of difference in LLIN effectiveness, we limited our assessment to a costing exercise. When the study LLINs were purchased (in 2019), the pyrethroid-pyriproxyfen LLINs (Royal Guard, $2.28 USD per net) were slightly less expensive than the pyrethroid-PBO LLINs (PermaNet 3.0, $2.51 USD per net). As of April 2023, market prices of the pyrethroid-pyriproxyfen LLINs were $0.77 USD lower per net than pyrethroid-PBO LLINs ($2.30 USD for Royal Guard vs $3.07 USD for PermaNet 3.0) [22].

## Discussion

In this pragmatic, cluster-randomised trial embedded into Uganda’s 2020–2021 LLIN distribution campaign, we compared the impact of two next generation LLINs on malaria incidence across 32 districts in Uganda. Despite a marked reduction in malaria incidence following LLIN distribution in both study arms, there was no difference in malaria incidence among residents of all ages, parasite prevalence among children 2-10 years of age, or anaemia prevalence among children 2-4 years of age from clusters that received pyrethroid-pyriproxyfen LLINs compared to those that received pyrethroid-PBO LLINs. Given the lack of difference in effectiveness of the two LLINs studied, we did not conduct a cost-effectiveness analysis as originally planned. Faced with similar effectiveness, the choice of LLIN should be guided by price and other factors. Importantly, LLINs will only be effective if coverage is high, and nets are used. Two years post-distribution, we found only around 40% of households were adequately covered by LLINs, while just over 60% of residents reported sleeping under a LLIN the previous night.

Pyrethroid-PBO LLINs were superior to pyrethroid-only LLINs in two trials conducted in east Africa, including the original LLINEUP trial in Uganda [13–15]. A Cochrane systematic review incorporating both trials concluded that pyrethroid-PBO LLINs reduced parasite prevalence compared to pyrethroid-only LLINs up to 21–25 months post-distribution (OR 0.79 [95% CI 0.67–0.95]), in areas with highly pyrethroid-resistant mosquitoes (defined as < 30% mosquito mortality in discriminating dose assays) [12]. Based on this evidence, the WHO issued a conditional recommendation in 2022 supporting the deployment of pyrethroid-PBO LLINs instead of pyrethroid-only LLINs ‘in areas with ongoing malaria transmission where the principal malaria vector(s) exhibit pyrethroid resistance’ [16]. However, pyrethroid-PBO LLINs have several potential limitations. Widespread pyrethroid resistance has been reported in Uganda [23, 24], and across Africa, with both knock-down and metabolic resistance described across several settings [11]. PBO is a synergist, not an insecticide, and can only restore sensitivity to pyrethroid insecticides if resistance is due to specific metabolic mechanisms. Moreover, PBO may fail to overcome high-level pyrethroid resistance [25]. Questions about the bioefficacy, durability, and lifespan of PBO-pyrethroid LLINs have also been raised [12, 16], given concerns about the rapid degradation of PBO from nets raised by studies conducted in Kenya and Tanzania [26, 27]. In Uganda, an evaluation of nets distributed in the first LLINEUP trial found that while the pyrethroid content of LLINs was relatively stable, the PBO content fell significantly over two years; by 55% in Olyset Plus and 58% in PermaNet 3.0 [17]. The bioefficacy of the pyrethroid-PBO nets also declined over time with only 46% and 79% of mosquitoes knocked down by Olyset Plus and PermaNet 3.0, respectively, after 25 months [17]. The WHO has highlighted the need for further research on the effectiveness of pyrethroid-PBO LLINs, specifically in areas of lower transmission intensity and where pyrethroid resistance is not driven by cytochrome P450 metabolism, as well as on the durability and cost-effectiveness of pyrethroid-PBO LLINs [16].

Treating LLINs with a combination of insecticides with different modes of action may improve efficacy and help to prevent or delay the spread of insecticide resistance. Pyriproxyfen is an insect growth regulator, which has traditionally been used as a larvicide [28], but also acts as a sterilising agent, reducing the fecundity (egg laying), fertility (production of viable offspring), and longevity of adult mosquitoes [29]. Pyriproxyfen has a different mechanism of action than pyrethroids, is effective at very low concentrations, and is safe in humans [28]. In a step-wedge, cluster-randomised trial conducted in Burkina Faso, pyrethroid-pyriproxyfen LLINs (Olyset Duo) were associated with lower clinical incidence in children aged 6–59 months, and lower entomological inoculation rates than pyrethroid-only LLINs (Olyset Net) [18]. More recently, a cluster-randomised trial conducted in Tanzania evaluated three next generation LLINs, including LLINs treated with pyrethroid-pyriproxyfen, pyrethroid-PBO, and pyrethroid-chlorfenapyr (a novel pyrrole insecticide), comparing their effect on parasite prevalence measured by RDT in children aged 6 months to 14 years, as compared to pyrethroid-only LLINs [19]. In this trial, parasite prevalence at 24 months post-distribution in the pyrethroid-pyriproxyfen LLIN arm was no different than in the pyrethroid-PBO control arm (37.5% vs 45.8%, adjusted OR 0.79 [95% CI 0.54–1.17]). Similarly, a cluster-randomised trial conducted in Benin studied pyrethroid-pyriproxyfen and pyrethroid-chlorfenapyr LLINs, as compared to pyrethroid-only LLINs, evaluating their effect on malaria incidence in children aged 6 months to 10 years [30]. This study also found no difference between the pyrethroid-pyriproxyfen and pyrethroid-only LLINs over 2 years following distribution (0.84 vs 1.03 malaria cases per child-year, hazard ratio 0.86 [95% CI 0.65-1.14]). In both trials, only the pyrethroid-chlorfenapyr LLINs were superior to the pyrethroid-only LLINs after two years [19, 30].

In March 2023, the WHO updated the guidelines on use of LLINs, supported by the recent results of the trials in Tanzania and Benin, offering a conditional recommendation for the deployment of pyrethroid-pyriproxyfen LLINs instead of pyrethroid-only LLINs in areas with pyrethroid resistance, and highlighting concerns about the cost-effectiveness of the pyrethroid-pyriproxyfen LLINs [16]. In addition, the WHO issued a conditional recommendation against the deployment of pyrethroid-pyriproxyfen LLINs instead of pyrethroid-PBO LLINs, noting that the evidence favours pyrethroid-PBO LLINs which were also deemed more cost-effective than pyriproxyfen LLINs. The LLINEUP2 trial adds to the evidence of the effectiveness of pyrethroid-pyriproxyfen LLINs, but calls into question the recent WHO recommendations, given that the pyrethroid-pyriproxyfen LLINs (Royal Guard, Disease Control Technologies, USA) were less expensive than pyrethroid-PBO (PermaNet 3.0, Vestergaard Sàrl, Switzerland) LLINs in our trial ($2.30 USD for Royal Guard vs $3.07 USD for PermaNet 3.0) [22], with similar effectiveness. LLIN prices fluctuate, however, so this finding should not be generalised; whichever net has a lower market price on a given day will be the most cost-effective.

Malaria incidence was the primary outcome measure of the LLINEUP2 trial, which was a strength of the study. Incidence of malaria, defined as the number of symptomatic cases of malaria occurring in a population at risk over time, is considered the gold standard for assessing malaria burden. However, cluster-randomised trials aiming to measure malaria incidence can be very expensive and logistically challenging. In this trial, we leveraged a platform of enhanced health facility-based malaria surveillance, which was built and supported by the Ugandan Malaria Surveillance Project [31]. Although routinely collected health facility-based data are often limited by incomplete reporting and lack of laboratory-confirmation, our enhanced health facility-based surveillance system acts as an intervention, improving data quality, ensuring nearly all cases of malaria are laboratory-confirmed, and improving case management. Moreover, by defining target areas around MRCs, collecting data on village of residence of malaria, and enumerating the target area populations to determine denominators, we estimated longitudinal measures of malaria incidence at an unprecedented scale across Uganda, at relatively low cost. Embedding large-scale, cluster-randomised trials into Uganda’s national LLIN distribution campaigns has provided an innovative, robust method to effectively evaluate next generation LLINs at scale, an exercise which would otherwise be very costly. This design provides a new paradigm for trials evaluating the ‘real-world’ effectiveness of malaria control interventions.

This study had several limitations. First, due to resource limitations, entomologic surveillance and assessments of LLIN bioefficacy and durability were not conducted. Second, we were not able to survey the clusters prior to LLIN distribution, again due to lack of resources, and thus have no baseline measures of LLIN coverage, parasite prevalence or anaemia. However, MRC surveillance activities were ongoing for at least three months prior to LLIN distribution, providing baseline measures of malaria incidence for all clusters. Third, although no difference in prevalence of anaemia in children aged 2-4 years was observed between the pyrethroid-pyriproxyfen and pyrethroid-PBO arms at the 12- and 24-month timepoints, anaemia prevalence dropped markedly in both arms between the two surveys (pyrethroid-pyriproxyfen 34.0% to 19.0%; pyrethroid-PBO 36.2% to 19.7%). Reasons for this change are unclear but likely relate to variation around the threshold cut-off for anaemia (haemoglobin < 11 g/dL) and the complex aetiology of anaemia. Although malaria is a major cause of anaemia in endemic areas, other factors contribute including nutritional status, helminths and other infections, and health system factors.

Our results suggest that pyriproxyfen-pyrethroid LLINs could be an effective alternative to pyrethroid-PBO LLINs in Uganda. Rotation of insecticides, including deployment of LLINs treated with different insecticides, has been recommended as a strategy to manage insecticide resistance [32]. Supported by the recent trials in Tanzania and Benin, the WHO has also issued new guidelines on LLINs treated with chlorfenapyr, offering a strong recommendation for the deployment of pyrethroid-chlorfenapyr LLINs instead of pyrethroid-only LLINs, and a conditional recommendation for deployment of pyrethroid-chlorfenapyr LLINs instead of pyrethroid-PBO LLINs in areas where malaria vectors are resistant to pyrethroids. However, the evidence supporting these new recommendations is limited and further research on the effectiveness, durability and lifespan of next generation LLINs is needed. To address the continuing challenge of insecticide resistance, we are conducting another cluster-randomised trial (LLINEUP3) embedded with the 2023 universal coverage campaign in collaboration with the Ugandan MOH. This study is comparing the effectiveness of pyrethroid-chlorfenapyr LLINs versus pyrethroid-PBO LLINs in 24 clusters located in 20 districts in Uganda.

## ACKNOWLEDGEMENTS

We would like to thank Emmanuel Arinaitwe, James Kapisi, Chris Ebong, Bridget Nzarubara, Faith Kagoya, Emmanuel Bakashaba, Isiko Joseph, Paul Oketch, Benjamin Guma, Jolly Job Odongo, Daniel Ochuli, Geoffrey Omia, Leuben Patrick Okello, Geoff Lavoy, Peter Mutungi, Nicholas Wendo, Yasin Kisambira, Catherine Pitt and the administration of the Infectious Diseases Research Collaboration for all their contributions. We would also like to acknowledge and thank the members of the Uganda National Malaria Control Division and the Liverpool School of Tropical Medicine for logistical and other support rendered as we carried out the data collection and analysis. We are grateful to the district administration, health and political leadership teams for all their support and guidance during community engagement in the 32 districts of the study area.

## Data Availability

De-identified participant data and a data dictionary defining each field in the set will be made publicly available at the time of publication on the ClinEpiDB website

https://clinepidb.org

## AUTHORS’ CONTRIBUTIONS

SGS, GD, MRK, MJD, SG, CMS, JIN and JO conceived and designed the study. SGS, GD, MRK, JIN, SG, JFN and CMS developed the procedures and drafted the protocol. SG, JFN, MJN, KS and IN collected the data, with oversight from SGS and MRK. IN and GD managed the data. GD, SG, KS, and SGS analysed the data. JIN, AE, MJD and MRK advised on the analysis. SG, JFN, MJN, DG and SGS drafted the manuscript. All authors reviewed the manuscript, provided input and approved the final version for publication.

## DECLARATION OF INTERESTS

The authors declare that they have no competing interests.

## FUNDING STATEMENT

This project was funded primarily by the US National Institutes for health, with additional funding from the Bill and Melinda Gates Foundation. The content of the manuscript is solely the responsibility of the authors.

## DATA SHARING STATEMENT

De-identified participant data and a data dictionary defining each field in the set will be made publicly available at the time of publication on the ClinEpiDB website (https://clinepidb.org). A link to the statistical analysis for the primary outcome has also been provided on the ClinEpiDB website (https://clinepidb.org/ce/app/workspace/analyses/DS_624583e93e/5My0liA/visualizations/1fd2ff57-954e-4c58-bf39-ebb113cc5a70).

## Notes

### Competing Interest Statement

The authors have declared no competing interest.

### Clinical Trial

NCT04566510. Registered 28 September 2020, https://clinicaltrials.gov/ct2/show/NCT04566510

### Author Declarations

The research included in this trial was conducted under two study protocols. The Implementation Project Pilot Study covered enumeration of the target areas and estimation of malaria incidence using health facility surveillance data. This project was approved by the Makerere University School of Medicine Research & Ethics Committee (SOMREC, Ref 2019-087), the Ugandan National Council of Science and Technology (UNCST, Ref HS 2659), and the University of California, San Franciso Human Research Protection Program Institutional Review Board (UCSF Ref 19-27957). The LLINEUP2 trial protocol provided approval for the full trial, including the cross-sectional surveys, and was approved by the Makerere University School of Medicine Research & Ethics Committee (SOMREC Ref 2020-193), Ugandan National Council for Science and Technology (UNCST Ref HS1097ES), the University of California San Francisco Human Research Protection Program Institutional Review Board (UCSF Ref 20-31769), and the London School of Hygiene & Tropical Medicine Ethics Committee (LSHTM Ref 22615). Prior to enrolment into the clinical surveys, written informed consent was obtained from a parent or guardian for all children aged less than 18 years and the child provided assent if they were aged 8 years or older.

